# Index Case Testing in Mozambique: a descriptive analysis of 7 years of implementation

**DOI:** 10.1101/2024.11.20.24317602

**Authors:** Nely Honwana, Guita Amane, Miguelhete Lisboa, Lindsay Templin, Meghan Swor, Ana Paula Simbine, Ema Samussone, Aleny Couto

## Abstract

**Introduction:** In 2014, Mozambique introduced index case testing (ICT) to increase the identification of people living with HIV (PLHIV), including among hard-to-reach populations. The strategy elicits PLHIV’s sexual contacts and biological children for testing at the community and facility levels, improving testing efficiency in detecting HIV among people. We analyzed HIV program data to understand ICT contributions to case identification since 2017.

**Methods:** Routine aggregated program data from the President’s Emergency Plan for AIDS Relief (PEPFAR) Monitoring, Evaluation, and Reporting data were abstracted for 2017–2023. Test positivity was calculated as positive tests over the total number of tests conducted by modality. Data were analyzed by calendar year (January–December), sex, age group (<15, 15– 49, ≥50 years), and ICT setting (community-based, facility-based).

**Results:** During 2017–2023, overall test positivity for all modalities combined decreased from 5.7% (410,430/7,207,532) to 2.9% (297,012/10,190,365), and ICT increased by 92% (from 389,940 tests done in 2017 to 748,265 in 2023). The contribution of ICT to the total number of positive tests increased from 9.0% to 24.5% (37,076/410,430 to 72,622/297,012); among men tested, the contribution of ICT to positive tests increased from 10.2% to 29.9%; among children <15 years tested, from 20.3% to 35.3%; and among 15–49-year-olds tested, from 10.3% to 25.3%. Almost two-thirds (65.3%) of positive tests through ICT were community-based (276,356/ 423,166), with a testing positivity rate of 10.5% (276,356/2,632,862). The overall ICT test positivity rate was 12.0% (423,166/3,535,787) compared to the overall test positivity rate of 4.1% (2,373,985/57,909,176).

**Conclusion:** Over the 7-year period, HIV testing through ICT almost doubled, and one of every four people who tested positive for HIV was detected through ICT. ICT efficiency is demonstrated by a test positivity that is three times higher (12%) than the overall test positivity (4%), with particular efficiencies among hard-to-reach age and sex groups.

## Introduction

It is estimated that in 2023, 377,083 people living with HIV (PLHIV) in Mozambique were unaware of their infection, or 15.4% of the nation’s total estimated PLHIV^1^. The United Nations Global AIDS Strategy 2021-2026 goal is for 95% of PLHIV to be aware of their HIV status by 2025^2^. As Mozambique nears this goal, identifying the remaining PLHIV becomes increasingly challenging.

In 2014, index case testing (ICT) was introduced in Mozambique as one possible strategy to identify PLHIV by offering HIV testing services to people who may have been exposed via the index case. These direct contacts include sexual partners from the past 12 months, all needle-sharing partners, and PLHIV’s biological children under 15 years of age as defined by the Mozambique Ministry of Health (MISAU) HTS guidance. To reduce future transmission, MISAU offers prevention services to clients who test negative. ICT helps identify serodiscordant couples (one partner is living with HIV while the other is not) and individuals who already know their status but have not accessed care or dropped out of care for some time^3^.

MISAU, with support from the U.S. President’s Emergency Plan for AIDS Relief (PEPFAR) and other partners, currently offers ICT in all 11 provinces both at health facilities and in communities, via mobile testing. MISAU recommends utilizing ICT when clients are either newly diagnosed as HIV-positive, have a high viral load (>1,000 copies/mL), and/or have abandoned ART^3^.

While ICT has been implemented in the last seven years, information on the long-term contribution of ICT and/or best practices in the Mozambique context is scarce. This paper aims to describe the contribution of ICT to overall case finding between 2017 and 2023 and Mozambique’s progress towards UNAIDS’s first 95 goal.

## Methods

We reviewed HIV testing results between calendar years 2017-2023 reported to PEPFAR through PEPFAR’s District Health Information System 2 (DHIS2) and Data for Accountability Transparency and Impact Monitoring (DATIM). HIV testing results are reported quarterly using the disaggregation and modalities for HIV testing defined by the Monitoring, Evaluation and Reporting (MER) Indicator Reference Guide^4^.

For this report, PLHIV on treatment refers to the number of PLHIV who are currently receiving ART. HIV positivity rate is the number of individuals testing positive among all individuals tested during the reporting period^3^. ICT contribution refers to the contribution of ICT to the number of HIV positive tests reported during the period.

Using MER reported results, we describe the proportion of positive test results by several types of disaggregation including age (<15, 15–49, ≥50, and unknown), sex (male, female), modality (index testing as distinct from all other testing modalities, such as voluntary testing or provider-initiated testing) and by geography (by province). In addition, we describe the differences in percent positive yield between these sub-categories and how that yield has changed over time to show the contribution of ICT to overall case finding in Mozambique.^1^

## Results

From 2017 to 2023, the annual number of HIV tests conducted in Mozambique increased by 41%, from 7,207,532 to 10,190,365. Across the same period, the number of HIV positive test results decreased by 28%, from 410,430 to 297,012. HIV testing increased by 115% among males 25+ (648,935 to 1,393,717) and 71% among females 25+ (1,503,765 to 2,570,604) and decreased by 22% among children, <15 (1,316,274 to 1,028,008). HIV positive results increased by 3% among males 25+ (92,003 to 94,877) and decreased 18% among females 25+ (135,042 to 110,065). The largest decrease in number of people testing positive (excluding unknowns) was among children (50% [410,430 to 297,012]). Across provinces, the median change in HIV tests conducted was 35% (ranging from -22% [750,981 to 567,239] in Gaza Province to 129% [1,088,44 to 2,495,732] in Zambezia Province), and the median decrease in HIV positive tests was -25% (range = -57% [38,292 to 16,538] in Gaza Province to 13% [50,340 to 56,907] in Nampula Province) (Table 1).

**Table 1:** Change in Total Tests and Total Positive Tests between 2017 and 2023.

|  | All Tests |  |  | Positive Tests |  |  |
| --- | --- | --- | --- | --- | --- | --- |
|  | 2017 | 2023 | Percent Change | 2017 | 2023 | Percent Change |
|  | 7,207,532 | 10,190,365 | 41% | 410,430 | 297,012 | -28% |
| **Age/Sex** |  |  |  |  |  |  |
| <15 | 1,316,274 | 1,028,008 | -22% | 23,656 | 11,806 | -50% |
| 15-24 | 1,935,903 | 3,134,858 | 62% | 88,593 | 58,319 | -34% |
| 25+ Females | 1,503,765 | 2,570,604 | 71% | 135,042 | 110,065 | -18% |
| 25+ Males | 648,935 | 1,393,717 | 115% | 92,003 | 94,877 | 3% |
| Unknown Age | 1,802,655 | 2,063,178 | 14% | 71,136 | 21,945 | -69% |
| **Province** |  |  |  |  |  |  |
| _Military Mozambique | 77,144 | 94,889 | 23% | 5,588 | 5,402 | -3% |
| Cabo Delgado | 813,539 | 833,453 | 2% | 27,792 | 23,049 | -17% |
| Cidade De Maputo | 334,016 | 606,155 | 81% | 35,860 | 17,375 | -52% |
| Gaza | 750,981 | 587,911 | -22% | 38,292 | 16,538 | -57% |
| Inhambane | 503,002 | 546,831 | 9% | 24,183 | 13,640 | -44% |
| Manica | 706,667 | 593,348 | -16% | 35,812 | 21,844 | -39% |
| Maputo | 632,011 | 926,960 | 47% | 41,581 | 20,005 | -52% |
| Nampula | 1,017,796 | 1,668,675 | 64% | 50,340 | 56,907 | 13% |
| Niassa | 226,548 | 408,242 | 80% | 11,105 | 12,071 | 9% |
| Sofala | 534,004 | 731,383 | 37% | 43,411 | 30,154 | -31% |
| Tete | 523,380 | 696,786 | 33% | 21,243 | 19,344 | -9% |
| Zambezia | 1,088,444 | 2,495,732 | 129% | 75,223 | 60,683 | -19% |

From 2017 to 2023, the ICT contribution to overall tests conducted increased from 5% (389,940/7,207,532) to 7% (748,265/10,190,365) and the proportion of HIV positive tests through ICT increased from 9% (37,076/410,430) to 24% (72,677/297,012) (Graph 1). In 2023, the proportion of HIV positive tests through ICT among men 25+ was 31% (29,650/94,877) compared to 13% (12,397/92,003) in 2017. Similarly, ICT contribution to positive increased from 9% (12,705 to 135,042) to 25% (27,630/110,065) among women 25+. In 2023, the proportion of HIV positive tests (among those with known age) was lowest among adolescents (15% [9,038/58,319]) and highest among those aged <15 years (35% [4,174/11,806]) (See Table 2). The provincial median proportion of HIV positive tests in 2023 was 24% (ranging from 11% [2,211/20,005] in Maputo Province to 52% [10,078/19,344] in Tete Province). All provinces, except for Maputo Province, demonstrated an increase in ICT contribution to people testing positive and an increase in absolute number of people testing positive identified via ICT between 2017 and 2023 (Table 3).

**Table 2:**
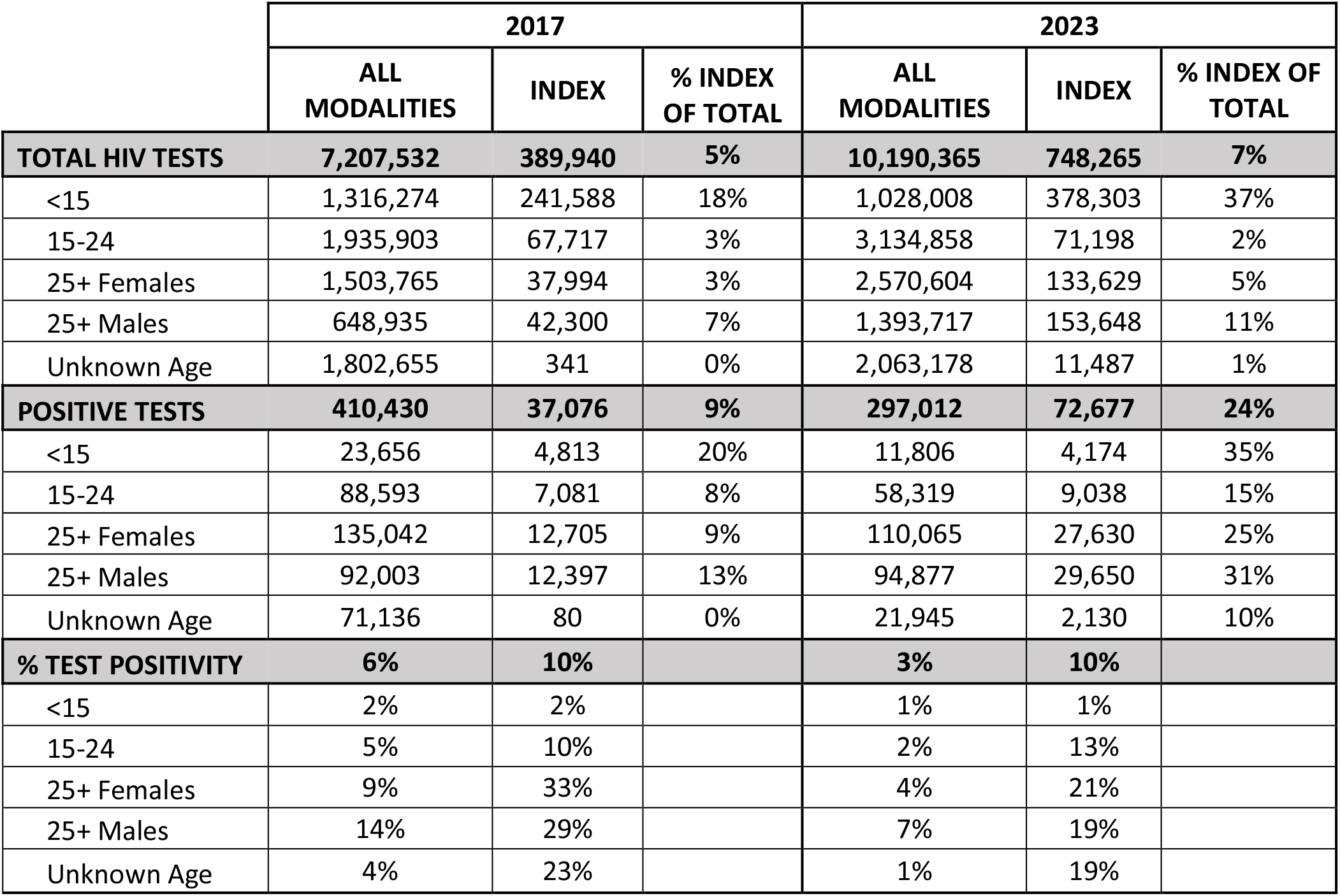
Percent Contribution of Index Testing by Modality and Age and Sex, 2017 and 2023.

**Table 3:** Percent Contribution of Index Testing by Modality and Province, 2017 and 2023.

|  | 2017 |  |  | 2023 |  |  |
| --- | --- | --- | --- | --- | --- | --- |
|  | ALL MODALITIES | INDEX | % INDEX OF TOTAL | ALL MODALITIES | INDEX | % INDEX OF TOTAL |
| **TOTAL HIV TESTS** | **7,130,388** | **388,086** | **5%** | **10,095,476** | **739,756** | **7%** |
| _Military Mozambique | 77,144 | 1,854 | 2% | 94,889 | 8,509 | 9% |
| Cabo Delgado | 813,539 | 60,324 | 7% | 833,453 | 33,121 | 4% |
| Cidade De Maputo | 334,016 | 36,100 | 11% | 606,155 | 48,225 | 8% |
| Gaza | 750,981 | 120,604 | 16% | 587,911 | 47,346 | 8% |
| Inhambane | 503,002 | 6,276 | 1% | 546,831 | 33,021 | 6% |
| Manica | 706,667 | 9,040 | 1% | 593,348 | 48,354 | 8% |
| Maputo | 632,011 | 81,761 | 13% | 926,960 | 60,143 | 6% |
| Nampula | 1,017,796 | 4,466 | 0% | 1,668,675 | 105,679 | 6% |
| Niassa | 226,548 | 4,665 | 2% | 408,242 | 21,304 | 5% |
| Sofala | 534,004 | 18,717 | 4% | 731,383 | 56,602 | 8% |
| Tete | 523,380 | 12,205 | 2% | 696,786 | 53,686 | 8% |
| Zambezia | 1,088,444 | 33,928 | 3% | 2,495,732 | 232,275 | 9% |
| **POSITIVE TESTS** | **404,842** | **36,547** | **9%** | **291,610** | **71,138** | **24%** |
| _Military Mozambique | 5,588 | 529 | 9% | 5,402 | 1,484 | 27% |
| Cabo Delgado | 27,792 | 852 | 3% | 23,049 | 4,024 | 17% |
| Cidade De Maputo | 35,860 | 6,351 | 18% | 17,375 | 4,383 | 25% |
| Gaza | 38,292 | 6,317 | 16% | 16,538 | 3,689 | 22% |
| Inhambane | 24,183 | 579 | 2% | 13,640 | 1,749 | 13% |
| Manica | 35,812 | 1,640 | 5% | 21,844 | 8,919 | 41% |
| Maputo | 41,581 | 9,320 | 22% | 20,005 | 2,211 | 11% |
| Nampula | 50,340 | 529 | 1% | 56,907 | 9,353 | 16% |
| Niassa | 11,105 | 677 | 6% | 12,071 | 2,967 | 25% |
| Sofala | 43,411 | 4,492 | 10% | 30,154 | 9,194 | 30% |
| Tete | 21,243 | 1,693 | 8% | 19,344 | 10,078 | 52% |
| Zambezia | 75,223 | 4,097 | 5% | 60,683 | 14,571 | 24% |
| **% TEST POSITIVITY** | **6%** | **9%** |  | **3%** | **10%** |  |
| _Military Mozambique | 7% | 29% |  | 6% | 17% |  |
| Cabo Delgado | 3% | 1% |  | 3% | 12% |  |
| Cidade De Maputo | 11% | 18% |  | 3% | 9% |  |
| Gaza | 5% | 5% |  | 3% | 8% |  |
| Inhambane | 5% | 9% |  | 2% | 5% |  |
| Manica | 5% | 18% |  | 4% | 18% |  |
| Maputo | 7% | 11% |  | 2% | 4% |  |
| Nampula | 5% | 12% |  | 3% | 9% |  |
| Niassa | 5% | 15% |  | 3% | 14% |  |
| Sofala | 8% | 24% |  | 4% | 16% |  |
| Tete | 4% | 14% |  | 3% | 19% |  |
| Zambezia | 7% | 12% |  | 2% | 6% |  |

From 2017 to 2023, the annual ICT test positivity rate remained the same at 10%, compared to the overall test positivity rate, which declined from 6% (410,430/7,207,532) to 3% (297,012/10,190,365). In 2023, the overall and ICT test positivity rates among males 25+ were 7% (94,877/1,393,717) and 19% (29,650/153,648), respectively. Among females 25+, the test positivity rates were 4% overall (110,065/2,570,604), and, via ICT, 21% (27,630/133,629). Positivity among children was 2% for both overall and ICT testing in 2017. As of 2023, this positivity has decreased to 1% for both ICT and all modalities (see Table 1). In 2023, the provincial median overall test positivity in 2023 was 3% (ranging from 2% [20,005/926,960] in Maputo Province to 6% [5,402/94,889] among military sites). The provincial median ICT test positivity in 2013 was 11% (ranging from 4% [2,211/60,143] in Maputo Province to 19% [10,078/53,686] in Tete Province) (see Table 3).

**Graph 1:**
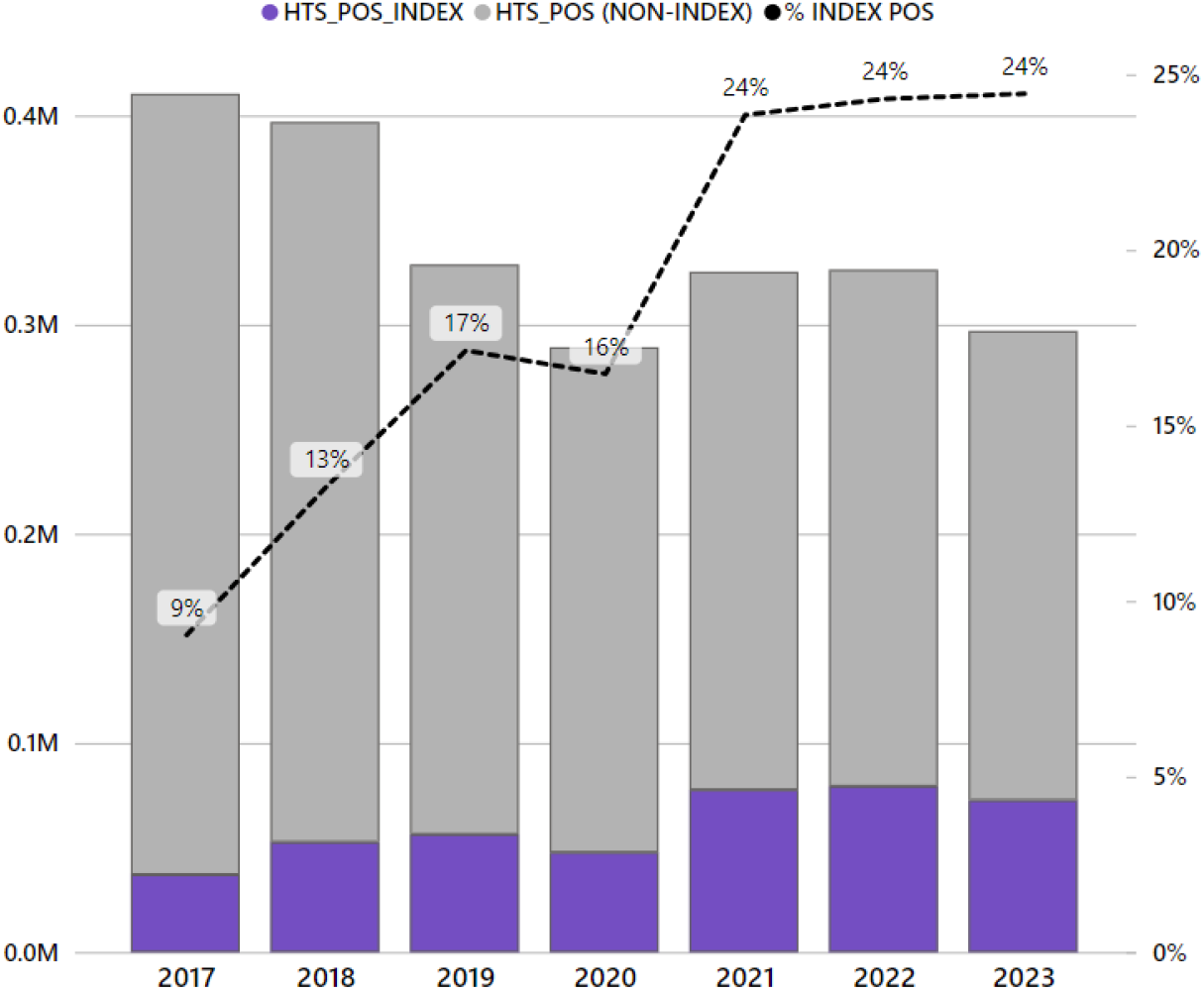
Case Identification and Index Positivity by Modality

## Discussion

From 2017 to 2023, the Mozambique Ministry of Health’s efforts to reach UNAIDS’ first 95% goal were aided by ICT, with a nearly two-fold increase in the proportion of HIV tests conducted via this testing modality. As the national test positivity rate from all HIV testing declined, the test positivity rate through ICT became responsible for nearly one-quarter of all cases identified, showing that this strategy will be increasingly useful as Mozambique nears 95% of PLHIV knowing their status. This is consistent with studies carried out in Gaza province, Mozambique, and Zambia, which show that ICT identifies PLHIV who are unaware of their HIV status, including hard-to-reach subgroups such as men^5,6^.

The identification of new PLHIV has slowed across every province and age group in Mozambique since 2017. However, PLHIV identified through ICT have either increased or slowed at a more gradual rate, showing the increased value of index testing to identify a shrinking population of PLHIV in Mozambique who remain unaware of their HIV status. Throughout the years of scale-up, including in 2023, the HIV test positivity rate was approximately 3 times higher through index testing compared to all modalities, therefore showing the efficiency of case finding via index testing. Because ICT helps identify sexual partners and biological children of PLHIV who are at a higher risk of similarly living with HIV, we expect to see higher HIV test positivity than through other modalities, as studied in Zimbabwe^7^.

For populations who infrequently visit health facilities and are relatively healthy, such as men and non-pregnant adolescents, index testing has proven to be an effective strategy in supporting case identification. For example, the number of positive cases identified among people 15-24 years old through index testing increased from 2017 to 2023, while the overall number of people testing positive identified through other testing modalities has decreased, demonstrating the critical importance of index testing. Our analysis showed that more men, non-pregnant adolescents, and young people were reached through ICT than other modalities.

Our analysis has limitations due to its reliance on partners reporting programmatic data. It was not possible to verify the testing modality reported or whether a person who reported positive was newly identified. In addition, the indicators for testing (regardless of HIV status) are not deduplicated at the patient level and, therefore, might represent some persons receiving multiple HIV tests per year.

## Conclusion

Between 2017 and 2023, while the proportion of HIV tests conducted through ICT increased 1.4-fold, the proportion of HIV positive tests via ICT increased 2.7-fold. As Mozambique continues to work towards the UNAIDS 95-95-95 goals, given its efficiency, ICT can continue to help identify the hard-to-reach PLHIV who are unaware of their status.

## Data Availability

All data produced are available from PEPFAR Monitoring, Evaluation, and Reporting program data.

https://data.pepfar.gov/datasets#PDD

## Funding Acknowledgement

This manuscript has been supported by the President’s Emergency Plan for AIDS Relief (PEPFAR) and through the Centers for Disease Control and Prevention (CDC). The findings and conclusions in this manuscript are those of the author(s) and do not necessarily represent the official position of the funding agencies (CDC, OGAC/PEPFAR).

## Footnotes

1 This activity was reviewed by CDC, deemed not research, and was conducted consistent with applicable federal law and CDC policy. See e.g., 45 C.F.R. part 46.102(I)(2), 21 C.F.R. part 56; 42 U.S.C. 241(d); 5 U.S.C. 552a; 44 U.S.C.c3501 et seq.

